# Prevalence and factors associated with stunting among children aged 6-59 months in Rubanda district, southwestern Uganda

**DOI:** 10.1101/2025.08.01.25332673

**Authors:** Ntabaare K. Rubahika, Timothy Nduhukire, Sam Tumwesigire, Agnes Napyo, James K. Tumwine

**Affiliations:** Department of Pediatrics and Child Health, Kabale University School of Medicine, Kabale, Uganda

**Author notes:** Correspondence: Ntabaare K. Rubahika. AOR: Adjusted Odds Ratio; CI: Confidence Interval.

**Keywords:** Stunting, Children, Southwestern Uganda, Rubanda, Poverty

## Abstract

**Background:** Childhood stunting remains a health and socioeconomic problem at global and national levels. Rubanda district, southwestern Uganda, contributed the highest cases of severe acute malnutrition seen at Kabale regional referral hospital.

**Objectives:** To determine the prevalence and factors associated with stunting among under-five children in Rubanda district, southwestern Uganda.

**Methods:** We used a cross-sectional study design and multistage cluster sampling to recruit 750 children. We took their anthropometric measurements and determined their stunting status. We also administered a questionnaire to the caregiver. STATA was used for descriptive and inferential analysis. Modified Poisson regression analysis was used to obtain crude and adjusted odds ratios.

**Results:** The prevalence of stunting was 52.27% (392/750). Factors associated with stunting included low wealth index (AOR 1.19, 1.01-1.39), male gender (AOR 1.15, 95% 1.01-1.31), age 24-35 months (AOR 1.27, 1.08-1.49), and delivery from private facility (AOR 0.74, 95% CI 0.58 - 0.94).

**Conclusion:** More than half of the children in Rubanda district, southwestern Uganda, were stunted. Factors associated included household poverty and being a male toddler. Alleviation of household poverty is essential to combat childhood stunting.

## INTRODUCTION

Globally, 21.3% of under-five children are stunted.(1) Whereas stunting is decreasing in other world regions, in Africa, it continues to rise.(1,2) Of note, Eastern Africa (1) contributes the highest proportion of 34.5%.

Although Uganda (3) has witnessed a drop in stunting from 30.7% to 26%, it is feared that with a low annual reduction rate,(4) it is unlikely to meet the Sustainable Development Goal target (5) of ending all forms of malnutrition by 2030. The southwestern region of Uganda, despite its high agricultural output, regional and community surveys have shown high levels of stunting: 41.5% from the Uganda Demographic Health Survey (UDHS) (3) and 41.1% by Kasajja et al.(6)

Stunting impairs skeletal growth (7) which largely passes unnoticed in communities where short stature is widespread.(8) In the short term, it increases the risk of death from infectious diseases,(9) and in the long term, it hinders the cognitive and economic achievement of an individual.(10,11) This, in turn, has an impact on the general societal socioeconomic situation.(7)

Rubanda district in southwestern Uganda was the leading contributor of children with severe acute malnutrition to Kabale regional referral hospital. The prevalence and factors associated with stunting in the district were unknown and so, we set out to fill this knowledge gap.

## MATERIALS AND METHODS

### Study design and setting

This was a cross-sectional community based study in Rubanda district located in the highland region of southwestern Uganda. The district had a population of 235,998 living in 480 villages. The study was carried out from 15^th^ October 2022 to 15^th^ February 2023.

### Study participants

All children aged 6-59 months whose caregivers gave informed consent were included in the study. Children who had not lived in the district for more than six months or who had stature limiting deformities such as scoliosis, cerebral palsy et cetera were excluded.

### Sample size

The sample size was calculated using the Kish Leslie formula for prevalence studies (12) denoted as N=p(1-p)z^2^/d^2^. We assumed a prevalence of 30.7% stunting based on UDHS figures from southwestern Uganda,(13) 95% confidence interval, and 5% precision, this gave a sample size of 327. We applied a design effect (14) of 2 and 10% non-response to increase the sample size to 720. Therefore, each of the 50 villages would contribute 14.4 (750/50) children. This approximated to 15 children per village, thus a final sample size of 750.

We used the Fleiss formula (15) in OpenEpi (16) to calculate the sample size for factors associated with stunting. Using the variable of age group 12-23 months (Odds ratio, 3.57) from a similar study in Ethiopia, (17) the sample size for associated factors was 440. Since 750 was the larger number, it was considered the overall sample size required to meet the study’s objectives. Selection of study participants

We used multi-stage cluster sampling to select the study participant. A total of 50 clusters were chosen from a list of villages in Rubanda district and each cluster contributed 15 children. To select the cluster, probability proportional to size sampling was used (18): A sampling interval was obtained by dividing the cumulative population by 50. We generated a random number that was less than or equal to the sampling interval. Cluster 1 was identified by locating the first village in which the cumulative population equaled or exceeded the random number. The second cluster was selected by adding the sampling interval to the first random number whereby the calculated new number fell at another point within the cumulative population. The consequent clusters were selected by repeating the process up to the 50^th^ cluster.

A household was defined as a group of people sharing the same kitchen.(18) The selection of the first household was determined randomly by the identification of an approximate geographical center in the village at which a pen was spun on the ground and wherever the tip of the pen pointed indicated the direction. The team then walked in the selected direction, counting the number of houses until the edge of the village. Then, a random number between one and the total number of houses along the directional line were selected and the house that corresponded to the number was the first house. The subsequent houses to be visited were chosen by proximity.

One eligible child was selected from each household. If there was more than one eligible child, the youngest was chosen to facilitate caregiver recall. Also, if the eligible children were from a multiple pregnancy, random sampling using the lottery method was employed to determine the study participant. If a selected household was without an eligible child, or consent was denied, or the eligible child was absent at the time of the visit, the most proximal household was visited.

### Study variables

The dependent variable was stunting. The independent variables were categorized as child, caregiver, and household characteristics. Child factors included age, sex, religion, ethnicity, antenatal care (ANC) attendance, type of pregnancy, place of delivery, birth weight, birth order, possession of child health card (CHC), vaccination status, growth monitoring status, distance to primary health care (PHC) facility, type of PHC facility, recent diarrhea, recent cough, recent fever, timely initiation of breastfeeding, prelacteal feeding, minimum dietary diversity, and minimum meal frequency. Caregiver factors included caregiver relationship, mother’s age, mother’s marital status, mother’s education, mother’s occupation, mother’s nutritional status, mother’s pregnancy status, father’s nutritional status, father’s education, and father’s occupation. Household factors included family size, number of under-five children, wealth index, food security, hand hygiene, presence of toilet facility, and presence of a waste dump site.

### Data collection

We measured the child’s height/length using a portable stadiometer (Model number: S0114530, UNICEF, Sichuan, China) and the mother’s/father’s mid-upper arm circumference (MUAC) using an adult MUAC tape (Model number: S0145630, UNICEF, China) in accordance with Food and Nutrition Technical Assistance (FANTA) protocol.(19) To ensure accuracy, the measurements were taken twice by two research assistants and the average of the two readings taken. This information (height/length) together with the child’s date of birth was entered into the 2006 WHO Anthro 3.2.1 software which calculated the Z-score. If the height/length-for-age Z-score was below -2, the child was considered stunted. Finally, adults whose MUAC was less than 22 cm were considered wasted.

We reviewed the child’s health card (CHC) to collect information about the vaccination and growth monitoring status. If the CHC was absent, a picture showing vaccine injection sites was shown to the caregiver to aid recall. Growth monitoring was considered adequate if done once every three months.(20)

The distance to the primary health care (PHC) facility was determined by a corroboration of the village health team’s (VHT) report and the caregivers walking time to the facility. A walking time of between 45-60 minutes was the standard for a health facility within 5 Km.(21)

We did a 24-hour dietary recall and determined the child’s dietary diversity. We cross-checked the child’s diet against the WHO food groups namely (i) Grains, roots, and tubers, (ii) Legumes and nuts, (iii) Dairy products, (iv) Flesh foods, (v) Eggs, (vi) Vitamin A rich fruits and vegetables, and (vii) Other fruits and vegetables. The diet was considered adequate if he/she received a minimum of four or more food groups. We asked the number of times the child had a main course meal to determine the meal frequency. Having three or more meals was considered adequate.

We collected information about household income, ownership of livestock, area of residence (urban or rural), ownership of house of residence, ownership of a financial account, ownership of agricultural land, ownership of non-agricultural land, and possession of piped water and used it to construct an asset score using Principal Component Analysis (PCA).(3) The score was categorized into high, medium, and low wealth indexes.

We evaluated food accessibility using the standardized “Household Food Insecurity Access Scale” questionnaire developed by FANTA project.(22) The caregivers were asked questions that sought to evaluate anxiety and uncertainty about food access, insufficient food quality, and insufficient food intake within the 30 days preceding the survey. From these questions, we constructed a binary outcome of food security or food insecurity.

An interviewer administered questionnaire was administered to collect and record information on the above and other variables.

### Data analysis

We entered data using EPI data 3.1, exported it to, and analyzed it in STATA version 17.0. Descriptive analyses were used to describe the percentages and frequency of the variables in the study. The prevalence of stunting was estimated and the 95% confidence interval determined. We used modified Poisson regression analysis for bivariable and multivariable analysis. All independent variables that were associated with stunting with a p-value <0.20 at bivariable analysis were exported to multivariable analysis. At multivariable analysis, a p-value of <0.05 was considered to be statistically significant.

### Quality control

A *Rukiga* language (the native language) expert did back-translation of the consent and data collection tool. The research assistants were registered nurses (diploma holders) proficient in *Rukiga* and who were trained before the field work.

### Ethical considerations

We obtained permission to carry out the study from Mbarara University of Science and Technology Research Ethics Committee: MUST-2022-493. Administrative clearance was obtained from the District Health Officer, Chief Administrative Officer, and Local Council Chairperson of each study village of Rubanda district. We obtained written informed consent from the mother/caretaker of the eligible child prior to the data collection process. Participant information was handled in confidence. Children and/or caregivers that were found to be with severe malnutrition or any other serious illness were linked to the nearest health facility for care. All the research procedures reported were done in accordance with the Declaration of Helsinki.

## RESULTS

### Baseline characteristics of study participants

Most of the study participants were 6 - 23 months of age, 332 (44.3%). The median age was 27.12 months (IQR, 15 - 38). Half the study participants (50.4%) were female. Only 10.8% had had adequate growth monitoring and half of the children had suffered from cough. The other characteristics are shown in Table 1.

**Table 1:** Child characteristics among children aged 6-59 months in Rubanda district.

| Variable | Total, N = 750<br>n | Percentage<br>% |
| --- | --- | --- |
| Age (months) |  |  |
| 6 – 23 | 332 | 44.3 |
| 24 - 35 | 199 | 26.5 |
| 36 – 59 | 219 | 29.2 |
| Sex |  |  |
| Female | 378 | 50.4 |
| Male | 372 | 49.6 |
| Religion |  |  |
| Christian | 741 | 98.8 |
| Muslim | 9 | 1.2 |
| Ethnicity |  |  |
| Mukiga | 742 | 98.9 |
| Others | 8 | 1.1 |
| ANC Attendance |  |  |
| Yes | 684 | 91.2 |
| Unknown | 66 | 8.8 |
| Type of Pregnancy |  |  |
| Single | 724 | 96.5 |
| Multiple | 26 | 3.5 |
| Place of delivery |  |  |
| Public Health Facility | 467 | 62.3 |
| Private Health Facility | 162 | 21.6 |
| Home | 59 | 7.9 |
| Unknown | 62 | 8.3 |
| Birth weight |  |  |
| High (> 3.9 Kg) | 57 | 7.6 |
| Low (< 2.5 Kg) | 47 | 6.3 |
| Unknown | 123 | 16.4 |
| Normal (2.5 – 3.9 Kg) | 523 | 69.7 |
| Birth order |  |  |
| > 4<sup>th</sup> | 158 | 21.1 |
| =< 4<sup>th</sup> | 592 | 78.9 |
| Physical possession of CHC |  |  |
| Yes | 708 | 94.4 |
| No | 42 | 5.6 |
| Adequate vaccination |  |  |
| Yes | 673 | 89.7 |
| No | 77 | 10.3 |
| Adequate growth monitoring |  |  |
| Yes | 81 | 10.8 |
| No | 669 | 89.2 |
| Distance to PHC facility |  |  |
| =< 5 Km | 659 | 87.9 |
| > 5 Km | 91 | 12.1 |
| Type of PHC facility |  |  |
| Public | 550 | 73.3 |
| Private (PNFP/PFP) | 200 | 26.7 |
| Recent diarrhea |  |  |
| No | 544 | 72.5 |
| Yes | 206 | 27.5 |
| Recent cough |  |  |
| No | 363 | 48.4 |
| Yes | 387 | 51.6 |
| Recent fever |  |  |
| No | 530 | 70.7 |
| Yes | 220 | 29.3 |
| Time to initiation of breastfeeding |  |  |
| Immediately | 594 | 79.2 |
| > 1 hr | 96 | 12.8 |
| Unknown | 60 | 8.0 |
| Prelacteal feeds |  |  |
| No | 513 | 68.4 |
| Yes | 177 | 23.6 |
| Unknown | 60 | 8.0 |
| Minimum Dietary Diversity |  |  |
| No (< 4 WHO foods) | 383 | 51.1 |
| Yes (>= 4 WHO foods) | 367 | 48.9 |
| Minimum Meal Frequency |  |  |
| No (< 3 meals/day) | 379 | 50.5 |
| Yes (>= 3 meals/day) | 371 | 49.5 |

The majority of the primary caregivers were mothers, 659 (87.9%). Most of the mothers were between 20 - 29 years, 327 (43.6%). The median age was 29.62 (IQR, 24 - 34). Many of the mothers were married, 642 (85.6%). The smallest proportion of households, 194 (25.9%), had a high wealth index. Most of the households were food insecure, 542 (72.3%). These and other characteristics are shown in Table 2.

**Table 2:**
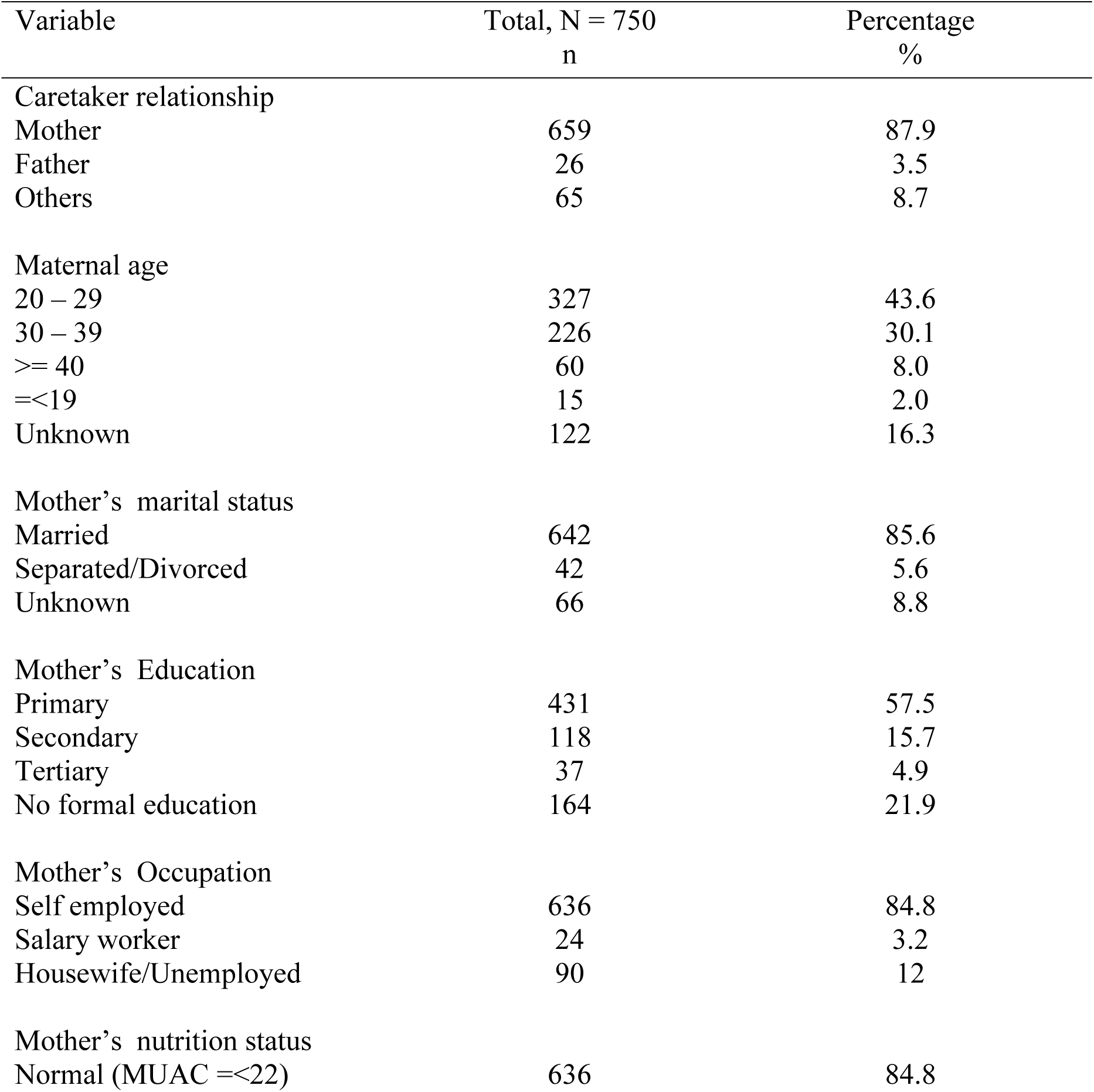
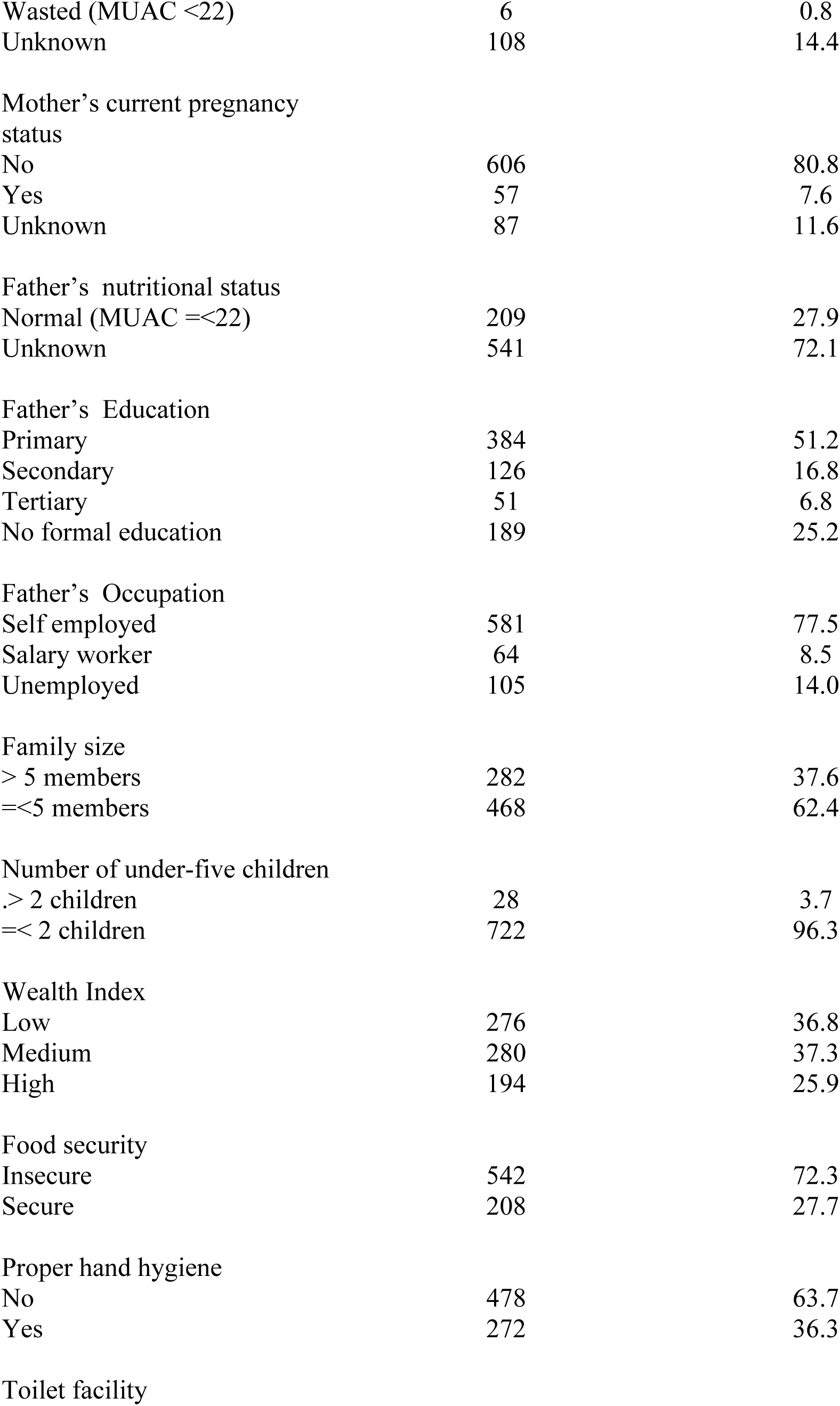
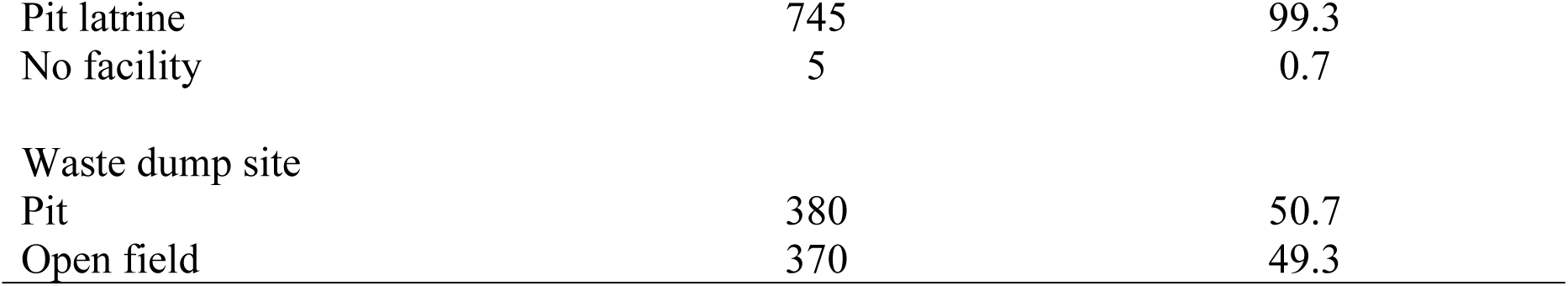
Caregiver and household characteristics among children aged 6-59 months in Rubanda district.

### Prevalence of stunting

Of the 750 children studied, 392 were stunted. The prevalence of stunting was 52.27% (95% CI, 48.62 - 55.89).

### Factors associated with stunting

Bivariable analysis was performed using modified Poisson analysis and the results are displayed in Table 3. The significant factors with a *p-*value <0.2 included: age, sex, ethnic group, place of delivery, birth weight, physical possession of CHC, distance to the PHC facility, type of PHC facility, recent fever, pre-lacteal feeds, Minimum Dietary Diversity (MDD), mother’s marital status, mother’s education, mother’s occupation, father’s education, father’s occupation, family size, wealth index, and food security.

**Table 3:** Factors associated with stunting among children aged 6-59 months in Rubanda district. (Bivariable modified Poisson analysis)

| Variable | Total<br>N=750<br>n(%) | Stunted<br>n(%) | Not<br>stunted<br>n(%) | Crude Odds<br>Ratio, 95% CI | p-value |
| --- | --- | --- | --- | --- | --- |
| Age (months) |  |  |  |  |  |
| 6 – 23 | 332 (44.3) | 161 (41.1) | 171 (47.8) | 1 |  |
| 24 - 35 | 199 (26.5) | 123 (31.4) | 76 (21.2) | 1.27 (1.07 – 1.51) | **0.006** |
| 36 - 59 | 219 (29.2) | 108 (27.6) | 111 (31.0) | 1.02 (0.81 – 1.28) | 0.885 |
| Sex |  |  |  |  |  |
| Female | 378 (50.4) | 183 (46.7) | 195 (54.5) | 1 |  |
| Male | 372 (49.6) | 209 (53.3) | 163 (45.5) | 1.16 (1.02 – 1.32) | **0.024** |
| Religion |  |  |  |  |  |
| Christian | 741 (98.8) | 387 (98.7) | 354 (98.9) | 1 |  |
| Muslim | 9 (1.2) | 5 (1.3) | 4 (1.1) | 1.06 (0.75 – 1.52) | 0.734 |
| Ethnicity |  |  |  |  |  |
| Mukiga | 742 (98.9) | 391 (99.7) | 351 (98.0) | 1 |  |
| Others | 8 (1.1) | 1 (0.26) | 7 (1.96) | 0.24 (0.45 – 1.25) | **0.09** |
| ANC Attendance |  |  |  |  |  |
| Yes | 684 (91.2) | 361 (92.1) | 323 (90.2) | 1 |  |
| Unknown | 66 (8.8) | 31 (7.9) | 35 (9.8) | 0.89 (0.65 – 1.23) | 0.465 |
| Type of Pregnancy |  |  |  |  |  |
| Single | 724 (96.5) | 378 (96.4) | 346 (96.7) | 1 |  |
| Multiple | 26 (3.5) | 14 (3.57) | 12 (3.35) | 1.03 (0.72 – 1.48) | 0.866 |
| Place of delivery |  |  |  |  |  |
| Public Health Facility | 467 (62.3) | 257 (65.6) | 210 (58.7) | 1 |  |
| Private Health Facility | 162 (21.6) | 66 (16.8) | 96 (26.8) | 0.74 (0.59 – 0.93) | **0.009** |
| Home | 59 (7.9) | 40 (10.2) | 19 (5.3) | 1.23 (1.01 – 1.50) | **0.039** |
| Unknown | 62 (8.3) | 29 (7.4) | 33 (9.2) | 0.85 (0.61 – 1.18) | 0.332 |
| Birth Weight |  |  |  |  |  |
| High (> 3.9 Kg) | 57 (7.6) | 28 (7.1) | 29 (8.1) | 1 |  |
| Low (< 2.5 Kg) | 47 (6.3) | 31 (7.9) | 16 (4.5) | 1.34 (0.91 – 1.99) | **0.142** |
| Unknown | 123 (16.4) | 67 (17.1) | 56 (15.6) | 1.11 (0.78 – 1.58) | 0.565 |
| Normal (2.5 – 3.9) | 523 (69.7) | 266 (67.9) | 257 (71.8) | 1.04 (0.74 – 1.44) | 0.838 |
| Birth order |  |  |  |  |  |
| > 4th | 158 (21.1) | 85 (21.7) | 73 (20.4) | 1 |  |
| =< 4th | 592 (78.9) | 307 (78.3) | 285 (79.6) | 0.96 (0.81 – 1.15) | 0.678 |
| Physical possession of CHC |  |  |  |  |  |
| Yes | 708 (94.4) | 374 (95.4) | 334 (93.3) | 1 |  |
| No | 42 (5.6) | 18 (4.6) | 24 (6.7) | 0.81 (0.59 – 1.11) | **0.189** |
| Adequate vaccination |  |  |  |  |  |
| Yes | 673 (89.7) | 355 (90.6) | 318 (88.8) | 1 |  |
| No | 77 (10.3) | 37 (9.4) | 40 (11.2) | 0.91 (0.68 – 1.22) | 0.534 |
| Adequate Growth Monitoring |  |  |  |  |  |
| Yes | 81 (10.8) | 41 (10.5) | 40 (11.2) | 1 |  |
| No | 669 (89.2) | 351 (89.5) | 318 (88.8) | 1.04 (0.79 – 1.35) | 0.793 |
| Distance to PHC facility |  |  |  |  |  |
| =< 5 Km | 659 (87.9) | 338 (86.2) | 321 (89.7) | 1 |  |
| > 5 Km | 91 (12.1) | 54 (13.8) | 37 (10.3) | 1.16 (0.97 – 1.38) | **0.105** |
| Type of PHC facility |  |  |  |  |  |
| Public | 550 (73.3) | 298 (76.0) | 252 (70.4) | 1 |  |
| Private | 200 (26.7) | 94 (24.0) | 106 (29.6) | 0.87 (0.72 – 1.05) | **0.142** |
| Recent diarrhea |  |  |  |  |  |
| No | 544 (72.5) | 285 (72.7) | 259 (72.4) | 1 |  |
| Yes | 206 (27.5) | 107 (27.3) | 99 (27.7) | 0.52 (0.48 – 0.57) | 0.91 |
| Recent cough |  |  |  |  |  |
| No | 363 (48.4) | 189 (48.2) | 174 (48.6) | 1 |  |
| Yes | 387 (51.6) | 203 (51.79) | 184 (51.4) | 1.01 (0.86 – 1.18) | 0.927 |
| Recent fever |  |  |  |  |  |
| No | 530 (70.7) | 268 (68.4) | 262 (73.2) | 1 |  |
| Yes | 220 (29.3) | 124 (31.6) | 96 (26.8) | 1.11 (0.96 – 1.30) | **0.164** |
| Time to initiation of Breastfeeding |  |  |  |  |  |
| Immediately | 594 (79.2) | 308 (78.6) | 286 (79.9) | 1 |  |
| > 1 hr | 96 (12.8) | 55 (14.0) | 41 (11.5) | 1.10 (0.91 – 1.35) | 0.320 |
| Unknown | 60 (8.0) | 29 (7.4) | 31 (8.7) | 0.93 (0.68 – 1.27) | 0.660 |
| Prelacteal Feeds |  |  |  |  |  |
| No | 513 (68.4) | 256 (65.3) | 257 (71.8) | 1 |  |
| Yes | 177 (23.6) | 107 (27.3) | 70 (19.6) | 1.21 (1.01 – 1.45) | **0.034** |
| Unknown | 60 (8.0) | 29 (7.4) | 31 (8.7) | 0.97 (0.69 – 1.35) | 0.851 |
| Minimum Dietary Diversity |  |  |  |  |  |
| No (< 4 WHO foods) | 383 (51.1) | 212 (54.1) | 171 (47.8) | 1 |  |
| Yes (>= 4 WHO foods) | 367 (48.9) | 180 (45.9) | 187 (52.2) | 0.89 (0.77 – 1.02) | **0.096** |
| Minimum Meal Frequency |  |  |  |  |  |
| No | 379 (50.5) | 192 (49.0) | 187 (52.2) | 1 |  |
| Yes | 371 (49.5) | 200 (51.0) | 171 (47.8) | 1.06 (0.92 – 1.23) | 0.405 |
| Caretaker relationship |  |  |  |  |  |
| Mother | 659 (87.9) | 348 (88.9) | 311 (86.9) | 1 |  |
| Father | 26 (3.5) | 13 (3.3) | 13 (3.6) | 0.95 (0.58 – 1.54) | 0.825 |
| Others | 65 (8.7) | 31 (7.9) | 34 (9.5) | 0.90 (0.66 – 1.24) | 0.529 |
| Mother's marital status |  |  |  |  |  |
| Married | 642 (85.6) | 343 (87.5) | 299 (83.5) | 1 |  |
| Separated/Divorced | 42 (5.6) | 18 (4.6) | 24 (6.70) | 0.80 (0.60 – 1.07) | **0.140** |
| Unknown | 66 (8.8) | 31 (7.9) | 35 (9.8) | 0.88 (0.64 – 1.21) | 0.426 |
| Mother's Education |  |  |  |  |  |
| Primary | 431 (57.5) | 241 (61.5) | 190 (53.1) | 1 |  |
| Secondary | 118 (15.7) | 55 (14.0) | 63 (17.6) | 0.83 (0.70 – 0.99) | **0.045** |
| Tertiary | 37 (4.9) | 10 (2.6) | 27 (7.5) | 0.48 (0.24 – 0.96) | **0.039** |
| No formal education | 164 (21.9) | 86 (21.9) | 78 (21.8) | 0.94 (0.77 – 1.14) | 0.510 |
| Mother's Occupation |  |  |  |  |  |
| Self employed | 636 (84.8) | 341 (87.0) | 295 (82.4) | 1 |  |
| Salary worker | 24 (3.2) | 8 (2.0) | 16 (4.5) | 0.62 (0.31 – 1.24) | **0.176** |
| Housewife/Unemployed | 90 (12) | 43 (11.0) | 47 (13.1) | 0.89 (0.72 – 1.10) | 0.290 |
| Mother's nutritional status |  |  |  |  |  |
| Normal | 636 (84.8) | 333 (85.0) | 303 (84.6) | 1 |  |
| Wasted | 6 (0.8) | 3 (0.8) | 3 (0.8) | 0.95 (0.38 – 2.40) | 0.922 |
| Unknown | 108 (14.4) | 56 (14.2) | 52 (14.5) | 0.99 (0.82 – 1.20) | 0.922 |
| Mother's current pregnancy status |  |  |  |  |  |
| No | 606 (80.8) | 319 (81.4) | 287 (80.2) | 1 |  |
| Yes | 57 (7.6) | 32 (8.2) | 25 (7.0) | 1.07 (0.85 – 1.35) | 0.587 |
| Unknown | 87 (11.6) | 41 (10.4) | 46 (12.8) | 0.89 (0.69 – 1.17) | 0.412 |
| Father's nutritional status |  |  |  |  |  |
| Yes | 209 (27.9) | 209 (27.9) | 108 (30.2) | 1 |  |
| Unknown | 541 (72.1) | 541 (72.1) | 250 (69.8) | 1.11 (0.94 – 1.32) | 0.218 |
| Father's Education |  |  |  |  |  |
| Primary | 384 (51.2) | 212 (54.1) | 172 (48.0) | 1 |  |
| Secondary | 126 (16.8) | 70 (17.9) | 56 (15.6) | 1.01 (0.85 – 1.19) | 0.941 |
| Tertiary | 51 (6.8) | 15 (3.8) | 36 (10.1) | 0.53 (0.35 – 0.81) | **0.003** |
| No formal education | 189 (25.2) | 95 (24.2) | 94 (26.3) | 0.91 (0.75 – 1.11) | 0.355 |
| Father's Occupation |  |  |  |  |  |
| Self employed | 581 (77.5) | 308 (78.6) | 273 (76.3) | 1 |  |
| Salary worker | 64 (8.5) | 26 (6.6) | 38 (10.6) | 0.77 (0.57 – 1.04) | **0.086** |
| Unemployed | 105 (14.0) | 58 (14.8) | 47 (13.1) | 1.04 (0.82 – 1.32) | 0.735 |
| Family size |  |  |  |  |  |
| > 5 members | 282 (37.6) | 160 (40.8) | 122 (34.1) | 1 |  |
| =<5 members | 468 (62.4) | 232 (59.2) | 236 (65.9) | 0.87 (0.77 – 0.99) | **0.039** |
| Number of under five children |  |  |  |  |  |
| > 2 children | 28 (3.7) | 18 (4.6) | 10 (2.8) | 1 |  |
| =< 2 children | 722 (96.3) | 374 (95.4) | 348 (97.2) | 0.81 (0.58 – 1.12) | 0.20 |
| Wealth Index |  |  |  |  |  |
| Low | 276 (36.8) | 154 (39.3) | 122 (34.1) | 1.35 (1.06 – 1.49) | **0.008** |
| Medium | 280 (37.3) | 152 (38.8) | 128 (35.8) | 1.03 (0.89 – 1.18) | 0.701 |
| High | 194 (25.9) | 86 (21.9) | 108 (30.2) | 1 |  |
| Food security |  |  |  |  |  |
| Insecure | 542 (72.3) | 305 (77.8) | 237 (66.2) | 1 |  |
| Secure | 208 (27.7) | 87 (22.19) | 121 (33.8) | 0.74 (0.59 – 0.93) | **0.009** |
| Proper hand hygiene |  |  |  |  |  |
| No | 478 (63.7) | 258 (65.8) | 220 (61.4) | 1 |  |
| Yes | 272 (36.3) | 134 (34.2) | 138 (38.6) | 0.91 (0.78 – 1.07) | 0.273 |
| Toilet facility |  |  |  |  |  |
| Pit-latrine | 745 (99.3) | 389 (99.2) | 356 (99.4) | 1 |  |
| No facility | 5 (0.7) | 3 (0.8) | 2 (0.6) | 1.15 (0.43 – 3.06) | 0.781 |
| Waste dump site |  |  |  |  |  |
| Pit | 380 (50.7) | 207 (52.8) | 173 (48.3) | 1 |  |
| Open field | 370 (49.3) | 185 (47.2) | 185 (51.7) | 0.92 (0.79 – 1.05) | 0.227 |

Multivariable analysis was done using modified Poisson regression analysis to determine factors independently associated with stunting and the results are displayed in Table 4. The independently associated factors included: age 24 – 35 months (AOR 1.27, 95% CI 1.08 – 1.49); male sex (AOR 1.15, 95% CI 1.01 – 1.31); delivery from a private health facility (AOR 0.74, 95% CI 0.58 – 0.94); and Low wealth index (AOR 1.19, 95% CI 1.39-1.01).

**Table 4:**
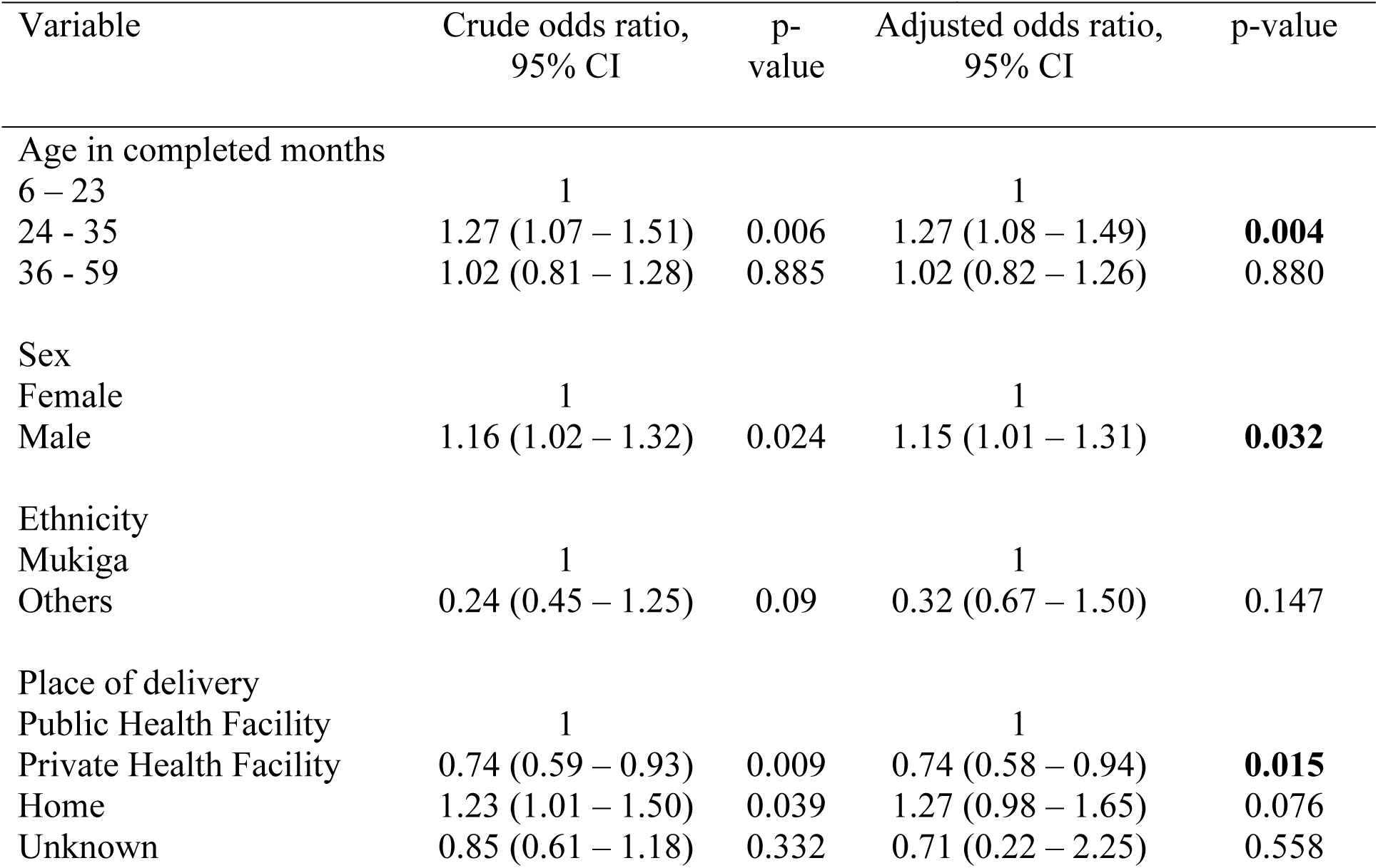
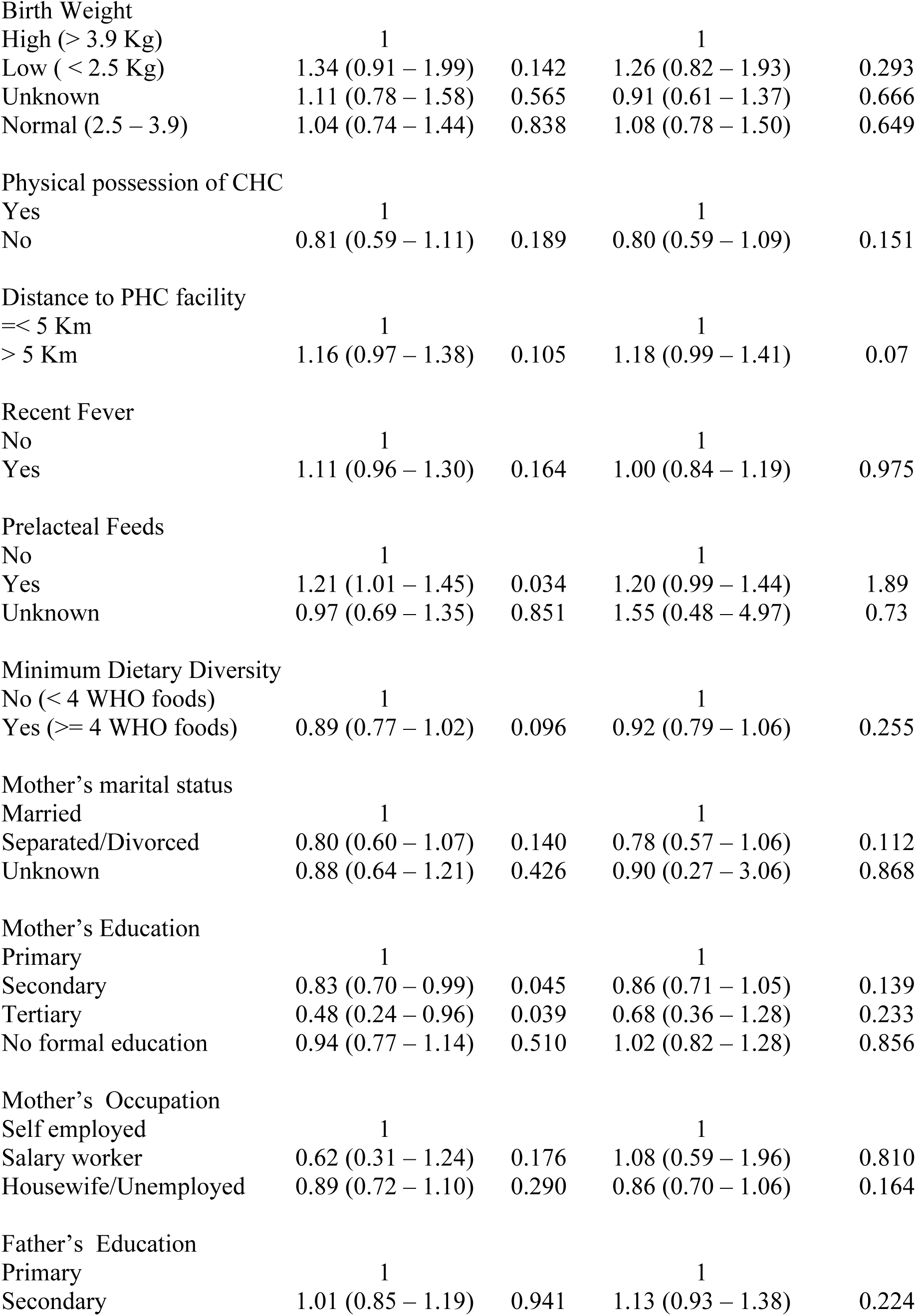
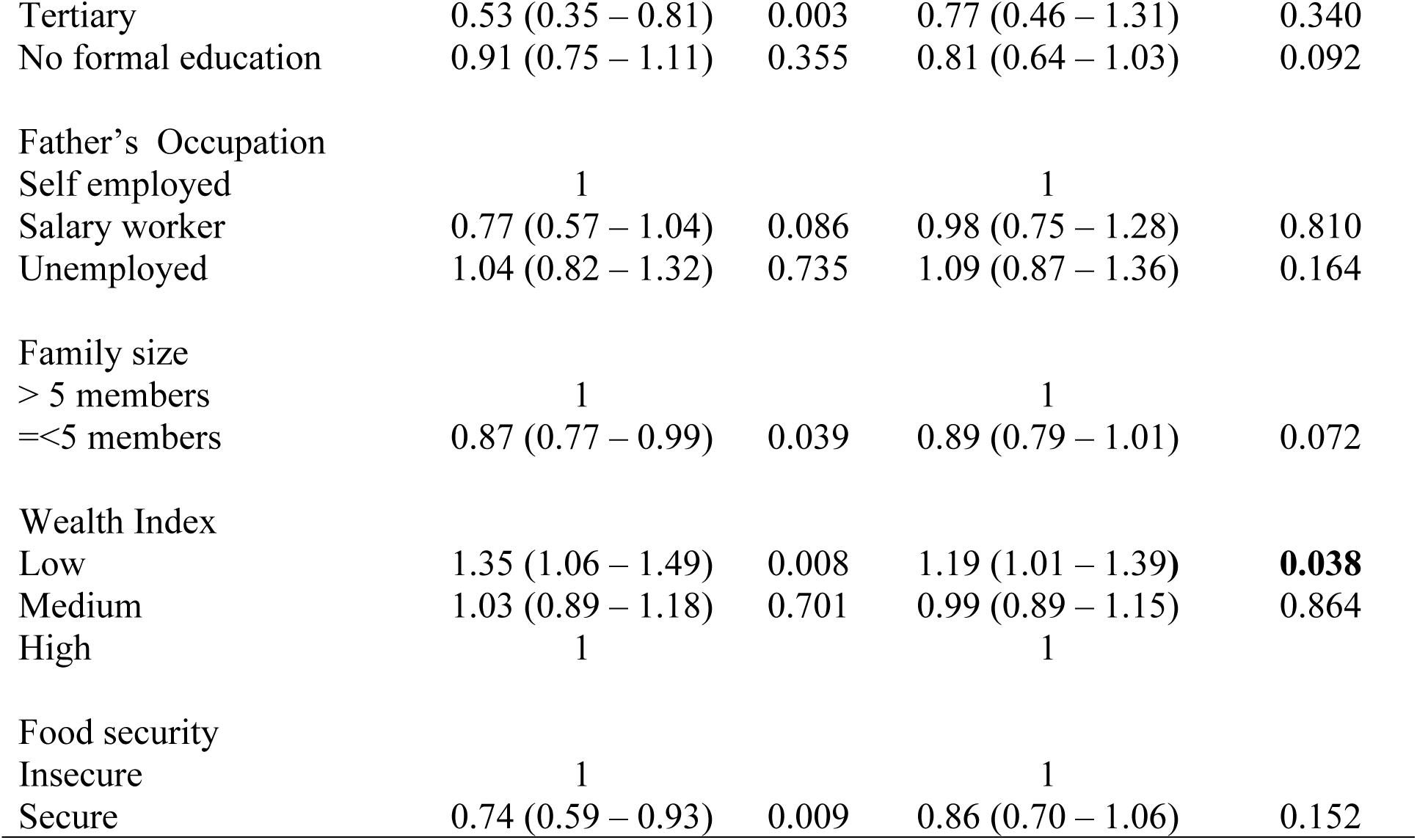
Factors that were independently associated with stunting among children aged 6 – 59 months in Rubanda district. (Multivariable modified Poisson analysis)

| Variable | Crude odds ratio,<br>95% CI | p-value | Adjusted odds ratio,<br>95% CI | p-value |
| --- | --- | --- | --- | --- |
| Age in completed months |  |  |  |  |
| 6 – 23 | 1 |  | 1 |  |
| 24 - 35 | 1.27 (1.07 – 1.51) | 0.006 | 1.27 (1.08 – 1.49) | **0.004** |
| 36 - 59 | 1.02 (0.81 – 1.28) | 0.885 | 1.02 (0.82 – 1.26) | 0.880 |
| Sex |  |  |  |  |
| Female | 1 |  | 1 |  |
| Male | 1.16 (1.02 – 1.32) | 0.024 | 1.15 (1.01 – 1.31) | **0.032** |
| Ethnicity |  |  |  |  |
| Mukiga | 1 |  | 1 |  |
| Others | 0.24 (0.45 – 1.25) | 0.09 | 0.32 (0.67 – 1.50) | 0.147 |
| Place of delivery |  |  |  |  |
| Public Health Facility | 1 |  | 1 |  |
| Private Health Facility | 0.74 (0.59 – 0.93) | 0.009 | 0.74 (0.58 – 0.94) | **0.015** |
| Home | 1.23 (1.01 – 1.50) | 0.039 | 1.27 (0.98 – 1.65) | 0.076 |
| Unknown | 0.85 (0.61 – 1.18) | 0.332 | 0.71 (0.22 – 2.25) | 0.558 |
| Birth Weight |  |  |  |  |
| High (> 3.9 Kg) | 1 |  | 1 |  |
| Low (< 2.5 Kg) | 1.34 (0.91 – 1.99) | 0.142 | 1.26 (0.82 – 1.93) | 0.293 |
| Unknown | 1.11 (0.78 – 1.58) | 0.565 | 0.91 (0.61 – 1.37) | 0.666 |
| Normal (2.5 – 3.9) | 1.04 (0.74 – 1.44) | 0.838 | 1.08 (0.78 – 1.50) | 0.649 |
| Physical possession of CHC |  |  |  |  |
| Yes | 1 |  | 1 |  |
| No | 0.81 (0.59 – 1.11) | 0.189 | 0.80 (0.59 – 1.09) | 0.151 |
| Distance to PHC facility |  |  |  |  |
| =< 5 Km | 1 |  | 1 |  |
| > 5 Km | 1.16 (0.97 – 1.38) | 0.105 | 1.18 (0.99 – 1.41) | 0.07 |
| Recent Fever |  |  |  |  |
| No | 1 |  | 1 |  |
| Yes | 1.11 (0.96 – 1.30) | 0.164 | 1.00 (0.84 – 1.19) | 0.975 |
| Prelacteal Feeds |  |  |  |  |
| No | 1 |  | 1 |  |
| Yes | 1.21 (1.01 – 1.45) | 0.034 | 1.20 (0.99 – 1.44) | 1.89 |
| Unknown | 0.97 (0.69 – 1.35) | 0.851 | 1.55 (0.48 – 4.97) | 0.73 |
| Minimum Dietary Diversity |  |  |  |  |
| No (< 4 WHO foods) | 1 |  | 1 |  |
| Yes (>= 4 WHO foods) | 0.89 (0.77 – 1.02) | 0.096 | 0.92 (0.79 – 1.06) | 0.255 |
| Mother's marital status |  |  |  |  |
| Married | 1 |  | 1 |  |
| Separated/Divorced | 0.80 (0.60 – 1.07) | 0.140 | 0.78 (0.57 – 1.06) | 0.112 |
| Unknown | 0.88 (0.64 – 1.21) | 0.426 | 0.90 (0.27 – 3.06) | 0.868 |
| Mother's Education |  |  |  |  |
| Primary | 1 |  | 1 |  |
| Secondary | 0.83 (0.70 – 0.99) | 0.045 | 0.86 (0.71 – 1.05) | 0.139 |
| Tertiary | 0.48 (0.24 – 0.96) | 0.039 | 0.68 (0.36 – 1.28) | 0.233 |
| No formal education | 0.94 (0.77 – 1.14) | 0.510 | 1.02 (0.82 – 1.28) | 0.856 |
| Mother's Occupation |  |  |  |  |
| Self employed | 1 |  | 1 |  |
| Salary worker | 0.62 (0.31 – 1.24) | 0.176 | 1.08 (0.59 – 1.96) | 0.810 |
| Housewife/Unemployed | 0.89 (0.72 – 1.10) | 0.290 | 0.86 (0.70 – 1.06) | 0.164 |
| Father's Education |  |  |  |  |
| Primary | 1 |  | 1 |  |
| Secondary | 1.01 (0.85 – 1.19) | 0.941 | 1.13 (0.93 – 1.38) | 0.224 |
| Tertiary | 0.53 (0.35 – 0.81) | 0.003 | 0.77 (0.46 – 1.31) | 0.340 |
| No formal education | 0.91 (0.75 – 1.11) | 0.355 | 0.81 (0.64 – 1.03) | 0.092 |
| Father's Occupation |  |  |  |  |
| Self employed | 1 |  | 1 |  |
| Salary worker | 0.77 (0.57 – 1.04) | 0.086 | 0.98 (0.75 – 1.28) | 0.810 |
| Unemployed | 1.04 (0.82 – 1.32) | 0.735 | 1.09 (0.87 – 1.36) | 0.164 |
| Family size |  |  |  |  |
| > 5 members | 1 |  | 1 |  |
| =<5 members | 0.87 (0.77 – 0.99) | 0.039 | 0.89 (0.79 – 1.01) | 0.072 |
| Wealth Index |  |  |  |  |
| Low | 1.35 (1.06 – 1.49) | 0.008 | 1.19 (1.01 – 1.39) | **0.038** |
| Medium | 1.03 (0.89 – 1.18) | 0.701 | 0.99 (0.89 – 1.15) | 0.864 |
| High | 1 |  | 1 |  |
| Food security |  |  |  |  |
| Insecure | 1 |  | 1 |  |
| Secure | 0.74 (0.59 – 0.93) | 0.009 | 0.86 (0.70 – 1.06) | 0.152 |

## DISCUSSION

The aim of this study was to determine the prevalence and factors associated with stunting among children aged 6 - 59 months in Rubanda district, southwestern Uganda.

### Prevalence of stunting

The prevalence of stunting was 52.27%. It is the highest magnitude of stunting documented in Uganda so far. This prevalence is twice the national average.(3) A recent study in Buhweju district in Uganda (23) reported a prevalence of 51%, while studies from Ethiopia (24,25) reported higher figures ranging from 59.9% to 64.5%.

### Factors associated with stunting

We found that children aged 24 - 35 months had a higher likelihood of being stunted. This has been found elsewhere.(26,27,28) However, our finding is different from that of Teferi et al. (29) who found that children 6 - 11 months were more likely to be stunted than their older counterparts. Stunting is an adaptive consequence of prolonged nutritional deprivation which might become evident as the child gets older.(8)

Boys were more likely to be stunted and this is corroborated by previous research.(30,31,32) However, two Ethiopian studies found the opposite.(33,34) It is thought that hormonal differences between sexes play a role, particularly luteinizing and follicle stimulating hormone.(35) Secondly, the differences in caring practices between male and female children has been implicated but this remains controversial.(36)

Being delivered from a private health facility was protective. It is possible that people who obtain care from private health facilities have a high wealth index which was found to be associated with less stunting in this study. Nevertheless, there is a dearth of literature associating delivery at private health facilities and stunting.

Children from poor households were likely to be stunted. Several studies (21,37,38) substantiate this finding. Poor families do not have the means to ensure food security and optimum dietary diversity (39) to meet the child’s nutritional and immunological needs.

### Strengths and limitations

Our study had some strengths. The sample size was large and participants were drawn from the entire district ensuring its representativeness. Also, we employed WHO Anthro software that enabled accurate determination of Z-scores. On the other hand, this being a cross-sectional study, causal relationship cannot be ascertained.

## CONCLUSION

Every second child aged 6 – 59 months in Rubanda district, southwestern Uganda was stunted. Boys and toddlers from poor households were likely to be stunted while children delivered from private facilities were protected. There is urgent need for interventions to combat childhood stunting.

## Data Availability

All relevant data are within the manuscript and its Supporting Information files.

## CONTRIBUTIONS

NKR, TN, ST, AN, and JKT were all involved in the conceptualization, data collection, analysis, and writing of the manuscript of this study.

## ACKNOWLEDGMENTS

We thank the children and their caregivers, research assistants, village health teams, and local government officials of Rubanda district for their invaluable involvement in the study.

